# The I-KAPCAM-AI-Q: A Novel Instrument for Evaluating Health Care Providers’ AI Awareness in Italy

**DOI:** 10.1101/2025.04.11.25325592

**Authors:** Vincenza Cofini, Laura Piccardi, Eugenio Benvenuti, Ginevra Di Pangrazio, Eleonora Cimino, Martina Mancinelli, Mario Muselli, Emiliano Petrucci, Giovanna Picchi, Patrizia Palermo, Loreta Tobia, Arcangelo Barbonetti, Giovambattista Desideri, Maurizio Guido, Franco Marinangeli, Leila Fabiani, Stefano Necozione

## Abstract

Background: Understanding healthcare providers’ readiness and attitudes is crucial for integrating AI in healthcare, yet no validated tool exists to evaluate these aspects among Italian physicians. This study developed and validated the Italian Knowledge, Attitudes, Practice, and Clinical Agreement between Medical Doctors and the Artificial Intelligence Questionnaire (I-KAPCAM-AI-Q). Methods: The validation process included expert review (n=18), face validity assessment (n=20), technical implementation testing, and pilot testing (n=203) with both residents and specialists. Results: The questionnaire demonstrated strong content validity (S-CVI/Ave=0.98) and acceptable internal consistency (Cronbach’s Alpha=0.7481, KR-21=0.832). Pilot testing revealed only 17% of participants had received digital technology training during medical education, while 91% showed clinical agreement with AI-proposed diagnoses. Knowledge in diagnostics was highest among AI applications (48%). Residents showed higher interest in technical support (58.3% vs 42.0%, p=0.021) and evidence-based validation (61.2% vs 47.0%, p=0.043) compared to specialists. Conclusion: The I-KAPCAM-AI-Q provides a reliable tool for assessing healthcare providers’ AI readiness and highlights the need for enhanced digital health education in medical curricula.

## Introduction

Since the advent of artificial intelligence (AI) in 1955, its applications have rapidly expanded within a dynamic digital landscape, driven by increasing public expectations influenced by social media, industry leaders, and healthcare professionals. Over the past decade, AI has effectively addressed numerous educational challenges, including language processing, reasoning, planning, and cognitive modelling [1]. Concerning medical education, AI can enhance it by recording teaching videos, facilitating distance learning, managing resources, and supporting virtual inquiry systems [2]. Additionally, AI can help process large repositories of medical data to enhance diagnostics and support problem-solving [3].

Generally speaking, AI in Medicine refers to the application of AI technologies, such as machine learning, deep learning, natural language processing, and other computational techniques, to improve healthcare outcomes, enhance clinical workflows, and support medical research. AI in medicine aims to leverage data-driven insights to aid decision-making, diagnosis, treatment planning, and personalized care [4]. Strong AI, which refers to the theoretical capability of a machine to comprehend or learn any intelligent task, can aid humans in addressing the challenges they encounter [5]. As AI continues to evolve, its capacity to enhance diagnostic precision and patient outcomes grows exponentially [6–7]. For instance, using AI-driven image recognition algorithms allows for achieving a 25% increase in diagnostic accuracy, compared to traditional manual methods, particularly in identifying early-stage anomalies in medical scans [8–9]. Nonetheless, a recent study revealed a significant lack of AI literacy among medical trainees: they often lack hands-on experience and a strong foundational knowledge of its application. Integrating AI into medical diagnostic workflows speeds up medical data analysis [10–12]. Undoubtedly, without the its implementation in the healthcare sector, according to the VALIDATE Project, many young medical doctors in Italy lack sufficient awareness and training in digital health and AI technologies, which are crucial for the digitalization of Italian healthcare system [13]. Indeed, AI allows systematic storage and management of medical data. Above all, the transition from paper-based records to electronic health records has vastly improved the accessibility and management of patient data. Electronic Health Records (EHRs) allow for the centralization of medical information, which can be quickly accessed by healthcare providers. In the meantime, Cloud Storage provides scalable and secure storage of large medical datasets, including images, genomic data, and patient records. A key contribution is also provided in data processing and categorization. For example, Natural Language Processing (NLP) is used to convert unstructured data (e.g., clinician’s handwritten notes or diagnostic reports) into structured, machine-readable data. On the other hand, Deep Learning and Image Recognition are applied to medical imaging to automatically identify anomalies (such as tumors) and categorize data for archiving; while Data Mining algorithms help identify patterns and trends across massive datasets, enhancing searchability and categorization of archived data [14].

Anyway, the lack of AI education represents a significant obstacle to effectively integrating AI-powered tools and technologies into clinical practice. Examining attitudes and perceptions of medical professionals, training physicians and AI proposals is essential for addressing this issue.

Growing concerns have arisen regarding the potential classification of AI as a separate legal entity, fueled by its ability to make autonomous decisions. The primary distinction between human and AI decision-making stems from the ethical implications of their actions [15]. Human beings’ decisions are profoundly shaped by moral considerations, an element entirely lacking in computer systems [16]. Undoubtedly, AI presents a remarkable opportunity for integrated home care services, effectively meeting the health needs of the elderly [17]. The Italian National Health Service prioritizes home care for elderly and non-self-sufficient patients for two compelling reasons. Firstly, it addresses the increasing demand for healthcare services due to an ageing population by delivering essential medical, rehabilitation and nursing care directly to patients’ homes. Secondly, providing care at homeit alleviates pressure on hospitals, leading to fewer emergency room visits and unnecessary hospitalizations, ultimately resulting in significant cost savings for the National Health System [18]. In 2021, approximately 3% of Italians aged 65 and older received home assistance amidst a total of 3 million individuals living with multiple chronic conditions and disabilities that necessitated continuous care [19]. This represents a significantly lower percentage when compared to other European nations, particularly those in Northern Europe. The Italian National Health Service recognizes the crucial need to promote the adoption of new Integrated Health Care systems as a primary objective. There is a growing global interest in the cutting-edge field of AI, particularly within public health, where efforts to enhance both lifespan and quality of life are increasingly prioritized. Moreover, the significance of patient telemonitoring —whether in medical facilities or at home— has become an even more urgent focus during the recent pandemic [20]. Italy is making significant strides toward transformative change with the welfare reform established in the National Recovery and Resilience Plan (PNRR), now officially enacted into law. Mission 6 of the PNRR aims to tackle regional disparities and improve the integration of health services across different care settings, focusing specifically on healthcare through telemedicine and AI innovation [21].

In the context of the Italian healthcare system, Clinical Decision support systems (CDSSs) commonly fall into two categories: diagnostic decision support and therapeutic decision support. For example, systems like Clinical Risk Prediction Models (CRPMs) are widely used to predict patient risk levels for various conditions, such as heart disease or sepsis [22]. Furthermore, systems like CPOE (Computerized Physician Order Entry), which are integrated with CDSS, can suggest appropriate drug prescriptions or dosages, ensuring both the safety and efficacy of treatments. In Italian hospitals, the adoption of these systems has been accelerating due to their potential to reduce medical errors and improve patient outcomes, despite the variability in their implementation across regions.[23].

Another integrated tool in healthcare is ChatGPT, a language model developed by OpenAI, primarily used for medical information retrieval, patient interaction, and clinical documentation assistance. While it does not directly make clinical decisions like CDSS, it serves as a conversational agent that can interact with healthcare professionals and patients to provide information, answer queries, or assist in decision-making workflows [24].

Nonetheless, a significant gap exists between AI training and its practical application, accompanied by considerable resistance among healthcare professionals to embrace AI technologies. For this reason, instruments to measure use, awareness, and knowledge of AI are mandatory.

Although some surveys are conducted worldwide to assess knowledge, attitudes, and the use of AI in medicine and educational medicine programs, a validated tool to measure these dimensions does not exist. To fill this gap, this study presents a comprehensive framework for designing and validating a questionnaire aimed at exploring the integration of AI in medical environments. It emphasizes the importance of understanding attitudes and clinical consensus between healthcare professionals and AI-driven recommendations. It will also assesses potential differences between residents and specialized medical doctors [25–28] to develop tailored educational programs and strategies to foster AI literacy and acceptance among medical students and residents.

This study aims to design and validate a comprehensive questionnaire (I-KAPCAM-AI-Q) that assesses the integration of AI technologies into clinical practice. Specifically, the I-KAPCAM-AI-Q focuses on the following issues:

1. Evaluating medical professionals’ knowledge and attitudes about AI in healthcare, focusing on their understanding of its capabilities, limitations, and impact on clinical workflows.
2. Investigating challenges and enablers of AI integration in clinical settings, analyzing aspects such as technology readiness, organizational support, regulatory concerns, and trust in AI systems.

Additionally, the second part of the I-KAPCAM-AI-Q evaluates how participants’ responses differ from ChatGPT’s solutions regarding diagnoses and interventions for real clinical scenarios previously submitted to the chatbot.

## Materials and methods

### Study Design and Setting

This study reports the validation process of a web-based questionnaire (I-KAPCAM-AI-Q) developed to assess Italian medical doctors’ perspectives on clinical AI and their clinical agreement for diagnosis and exams proposed by AI in clinical scenarios. Specifically, the clinical scenarios are based on real-world situations, developed by clinicians, and reviewed by experts to ensure their relevance. These scenarios were then submitted to ChatGPT [ChatGPT (OpenAI, 2024). ChatGPT (Version GPT-4). [AI language model]. https://openai.com]., and participants were asked to assess their agreement with the AI-generated diagnoses, which could include the correct diagnosis for comparison. The I-KAPCAM-AI-Q is important to assess the comprehension of AI’s role in diagnostic processes and to measure trust in AI-generated recommendations. Moreover, it provides current usage patterns of AI tools in clinical practice, allows identifying barriers and facilitators to AI adoption and measures willingness to incorporate AI into daily clinical workflows. This last point is crucial in adhering to the current Government guidelines regarding digital healthcare.

Before questionnaire development, we conducted a comprehensive literature review was conducted, searching articles from multiple databases, including PubMed, Scopus, and Web of Science. This review focused on physician and medical student acceptance of clinical AI, helping identify key themes and knowledge gaps in the field and informing the subsequent questionnaire development. In particular, the review focused on survey or cross-sectional studies with attention to the validation test reported [29–33].

Figure 1 shows the steps in developing the Italian Knowledge, Attitudes, Practice, and Clinical Agreement between Medical Doctors and the Artificial Intelligence Questionnaire (I-KAPCAM-AI-Q).

**Figure 1.**
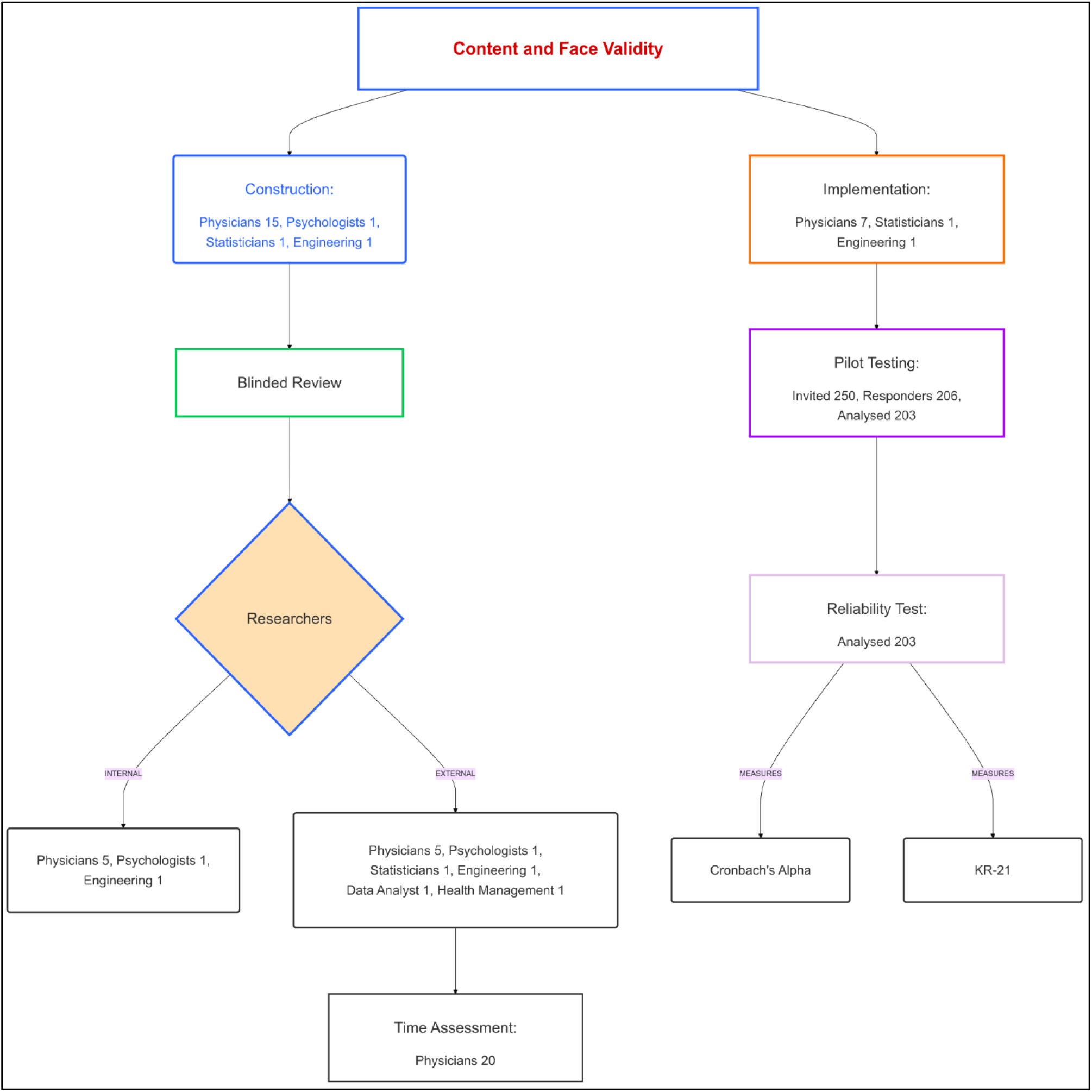
Validation Process Diagram.

### Participants

The validation process involved multiple groups of participants, as reported in Figure 1. All participants gave their consent to participate, and the subgroups consisted of the following:

- Three medical doctors (two residents in hygiene and preventive medicine and one practicing physician), one statistician, and one engineer for the initial design
- Ten specialized physicians representing diverse fields (hygiene and preventive medicine, endocrinology, infectious diseases, obstetrics and gynaecology, anesthesiology, occupational health, and geriatrics) for the development of clinical scenarios
- A panel of expert reviewers for content validation
- Volunteer physicians not involved in the research for time assessment
- Volunteer physicians not involved in the research for face validity assessment
- A sample of physicians for pilot testing, with analysis focusing on differences between specialization groups

### Validation Process

The validation methodology consisted of four sequential steps:

Step 1: Construction and Initial Validation Content validity

The questionnaire was organized into two parts to evaluate general perspectives and clinical reasoning comprehensively.

First Part: General Sections

The first part of the questionnaire consisted of six structured sections with items structured on answers yes/no or categorical or on a five-point Likert scale, and the last section for Open-ended Feedback:

1. Demographic and Work Information: this section collected data on respondents’ medical speciality, years of experience, and previous familiarity with digital technologies (Number of items: 9).
2. Knowledge about AI Technology in Medicine: this section assessed respondents’ knowledge of AI concepts and AI tools in clinical settings (Number of items: 3: 1-A question about participation in an AI course, designed to capture prior educational exposure; 2-A self-assessed level of knowledge question; - 3 A set of dichotomous questions to assess factual knowledge about specific AI concept plus 2 optional items linked to advanced knowledge declared).
3. AI Use and Experience in Medicine: this section focused on using AI technologies in clinical practice (Number of items: 3).
4. Attitudes and Awareness Regarding AI in Medicine: this section evaluated participants’ viewpoints on AI (Number of items: 7 on a five-point Likert scale).
5. Willingness to Use AI in Medicine: this section explored participants’ openness to integrating AI into their clinical workflows (Number of items: 3, with one item on a five-point Likert scale)
6. Willingness to Learn AI in Medicine: this section assessed participants’ interest in gaining knowledge and skills related to AI (Number of items: 2)
7. Open-ended Feedback: in this section, participants were invited to provide recommendations, suggestions, or general feedback about the questionnaire or the topics covered.

The validation content validity of the first part of the I-KAPCAM-AI-Q, was assessed through expert evaluation based on four key attributes: relevance, clarity, simplicity, and ambiguity. We utilized The Content Validity Index (CVI) was used to quantify the evaluation process, with items scoring above 0.79 considered sufficiently relevant [32]. Expert reviewers rated each item on a four-point scale from “not relevant” to “highly relevant”. Items scoring below the threshold were modified to improve clarity or alignment with study objectives. For face validity, volunteer physicians evaluated the questionnaire and were encouraged to seek clarification from the research team via telephone or email when needed. They documented concerns about specific questions while assessing response accuracy, comprehension of technical terminology, scenario realism, and completion time.

Second Part: Clinical Scenarios

The second part of the questionnaire included two categories of clinical scenarios: “universally applicable scenarios” and “specialty-specific scenarios,” aimed at evaluating the level of concordance between the respondents’ answers and ChatGPT’s diagnostic proposals.

The research team developed clinical scenarios and submitted them to AI for differential diagnoses and recommendations. All scenarios were based on real clinical cases with confirmed diagnoses and went through a structured validation process. Clinical experts from seven medical specialties reviewed the scenarios, considering clinical relevance (common presentations in clinical practice), appropriate complexity level, and ensuring clarity and completeness of the clinical information presented [34].

For each scenario, a structured presentation of the clinical case was provided to ChatGPT. The clinical scenarios were submitted to ChatGPT using the question: “What diagnosis would the patient receive based on the information provided?” [34]

The final tool included:

1. Universally Applicable Scenario: Upon completing the first general sections, all participants were presented with a clinical scenario designed to be universally applicable to medical graduates. This scenario was simple, ensuring that even newly qualified physicians with limited clinical experience could engage meaningfully. Its simplicity enabled consistent participation across varying levels of expertise, while providing valuable insights into general medical reasoning and decision-making skills.
2. Speciality-Specific Scenarios: The questionnaire also included seven distinct clinical scenarios tailored to the following specialities (Hygiene and Preventive Medicine, Infectious Diseases, Obstetrics and Gynecology, Anesthesiology, Geriatrics, Occupational Health, and Endocrinology).

The questionnaire was used to gather responses from participants to assess their level of agreement with AI’s proposals, using a 5-point Likert scale ranging from ‘strongly disagree’ to ‘strongly agree.’

The structure of the questions and the response formats were identical across all scenarios. All participants were asked the same question, and the response structure provided by ChatGPT was consistent for each scenario.

The main purpose of these scenarios was to provide context for specific constructs and behaviors, not to measure a latent variable, so psychometric validation was not considered necessary.

The initial validation study centered on analyzing the responses to the universally applicable scenario, which was created to be answered by all participants. Despite being included in the final validated tool, the specialty-specific scenarios were not analyzed during this phase, because the pilot sample had a small number of specialists per category.

At the end of the questionnaire, participants could report how they had the link to the questionnaire.

Step 2: Time Assessment and Questionnaire Implementation

Twenty expert readers (medical doctors) were enrolled to evaluate the time to complete the questionnaire. The time collected was related to the total time spent to fill all questionnaire items and the time spent in each section and clinical scenarios designed. This activity was also useful for verifying and validating participant data.

The Questionnaire was designed for future web-based research studies, and technical aspects were evaluated, including:

- Survey platform capabilities, such as adaptive skip logic implementation and real-time progress saving functionalities.
- Mobile device compatibility and cross-platform devices to ensure accessibility and usability
- Advanced data export functionalities for subsequent analysis
- User interface optimization, including dynamic progress indicators and clearly defined section transitions for enhanced respondent experience
- Integrated help text for the clarification of technical terminology, improving respondent comprehension.
- Error prevention mechanisms to mitigate data entry inconsistencies and enhance response accuracy.
- Comprehensive data management and security protocols, including encrypted data storage, GDPR-compliant handling [35], and secure export functionalities. Data management and security (data export functionality, encryption, GDPR-compliant data handling)
- Strict adherence to accessibility standards, ensuring full compliance with Web Content Accessibility Guidelines 2.1 (WCAG 2.1) [36] for inclusive digital survey deployment. Accessibility standards (full adherence to WCAG 2.1 guidelines)

Step 3: Pilot Testing Study

An online pilot test was conducted to investigate potential response differences between specific subgroups, particularly focusing on their medical specialization status. We analysed categorical variables using percentages and continuous variables using means and standard deviations. Group comparisons were performed using Chi-square, Fisher Exact, and Mann-Whitney U tests. Data were collected from July to September 2024.

Step 4: Reliability Assessment Internal consistency

For this step, Cronbach’s Alpha and Kuder-Richardson Coefficient (KR21) were used. Cronbach’s Alpha for Likert-scale items, particularly the eight items measuring opinions and willingness to use AI in medicine. Two items were reverse-coded for consistent directional interpretation. Values ≥0.90 were interpreted as excellent, 0.80-0.89 as good, and 0.70-0.79 as acceptable reliability [37]. Item-level statistics included item-test correlation, item-rest correlation, and the impact of item removal on Cronbach’s Alpha to evaluate individual item contributions to overall reliability. Kuder-Richardson Coefficient (KR21) was used for dichotomous data or all data split into categories. KR21 with values ≥0.80 indicates good to excellent reliability, and 0.70-0.79 is acceptable [38]. Internal consistency analysis was not performed for each dimension separately, as each dimension included both Likert-scale and dichotomous questions.

Analysis was performed using STATA 18 BE, setting alpha 0.05. Figure 1 was made using the Mermaid Diagramming and charting tool (https://mermaid.js.org/), and Figure 2 was from Jamovi 2.6.13.

**Figure 2.**
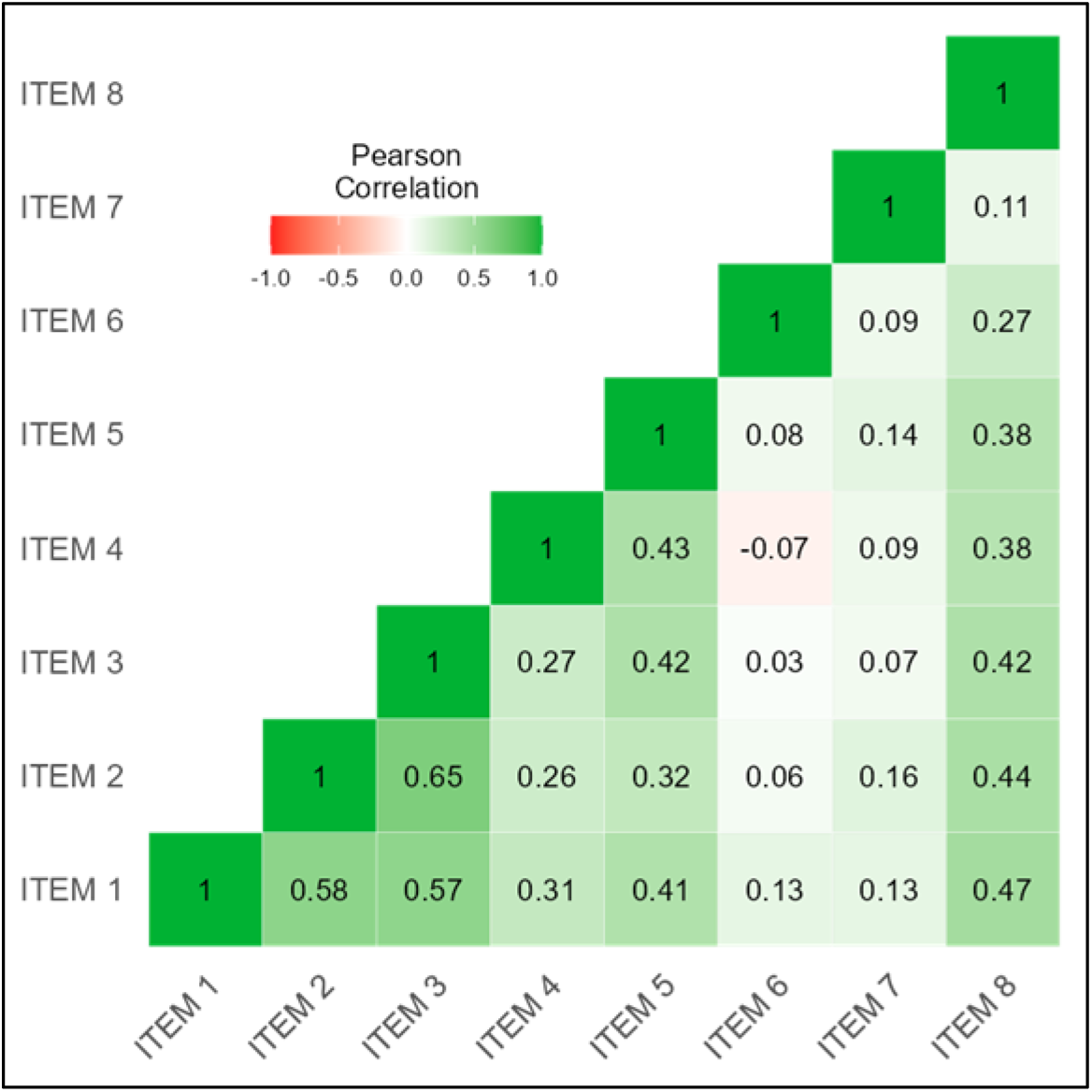
Correlation Heatmap.

## Results

The results presented in this section describe the validation process of the I-KAPCAM-AI-Q questionnaire through four sequential steps, each designed to evaluate a specific aspect of the instrument. The data collected during pilot testing provided interesting preliminary insights into AI perceptions in medicine. They were primarily used to verify the effectiveness and reliability of the assessment tool and should not be interpreted as representative of the general population of Italian physicians. The analysis of results therefore focuses on the psychometric properties of the questionnaire and its ability to detect significant differences between groups, thus demonstrating its utility as a research instrument

Step 1: The I-KAPCAM-AI-Q Validation

As reported in Figure 1, a panel of 18 experts from relevant medical or non-medical fields was convened to review the tool. For each item, I-CVI>0.88 S-CVI/Ave (Average Approach=0.98) and S-CVI/UA (Universal Agreement Approach=0.82). The content validity ratio (CVR) ranges from 0.78 to 1.

Step 2: Time Assessment and Implementation

To evaluate the practical feasibility and usability of the questionnaire, twenty physicians assessed the time required to complete the online questionnaire.

The average completion time for the questionnaire with the universal clinical scenario was 7.74 minuteswas 7.74 minutes (SD=3.65). The time spent completing the specialised clinical scenarios is reported in Table 1.

**Table 1.** - Time assessment to fill the questionnaire by medical speciality -.

| Speciality | Physicians | Meantime (Minutes) | Standard deviation (Minutes) |
| --- | --- | --- | --- |
| Hygiene and Preventive Medicine | 5 | 10.13 | 3.4 |
| Infectious Diseases | 2 | 11.91 | 10.3 |
| Obstetrics and Gynecology | 4 | 7.87 | 3.4 |
| Anesthesiology | 3 | 7.55 | 0.4 |
| Geriatrics | 5 | 10.76 | 7.8 |
| Occupational Health | 4 | 9.00 | 4.1 |
| Endocrinology | 5 | 6.19 | 1.1 |

Technical Implementation Results

After evaluating open-source survey platforms, LimeSurvey was selected as the optimal solution due to its comprehensive features and user-friendly interface [39–40]. The platform demonstrated capabilities aligned with our technical requirements, particularly in handling complex survey logic and ensuring data security compliance. System Implementation

The survey data collection was deployed on a dedicated hosting environment, integrating LimeSurvey with a high-performance LAMP stack (Linux [41], Apache [42], MySQL [43], PHP [44]). The system architecture was designed to ensure high availability, data integrity, and enhanced security measures through:

- Deployment of the latest stable LimeSurvey version
- MySQL was implemented as the database management system, supporting data storage and retrieval.
- Implementation of file and directory permission settings to strengthen system security and prevent unauthorized access.
- Configuration of survey parameters and interface language settings Implementation of user interface optimization, including dynamic progress indicators and clearly defined section transitions for enhanced respondent experience
- Implementation of industry-standard security protocols, including Transport Layer Security (TLS) encryption [45], Secure Sockets Layer (SSL) certification [46], and HyperText Transfer Protocol over Secure Socket Layer (HTTPS) [47] enforcement for secure data transmission

Technical Features and Functionality

The implemented system successfully demonstrated several key functionalities:

- Survey Logic and Data Management:
- Robust skip logic handling with accurate participant pathway direction
- Reliable progress-saving capability allowing session resumption
- Multi-format data export, including Comma-separated values (CSV) [48], Excel Open XML Spreadsheet (XLSX) [49]
- Secure data transmission and storage compliant with data protection regulations

Cross-Platform Compatibility:

- Verified functionality across web browsers (Google Chrome, Mozilla Firefox, Apple Safari)
- Responsive design for mobile devices (iOS and Android)
- Consistent display of all survey elements regardless of device

User Interface Elements:

- Dynamic progress indicators accurately reflecting completion status
- Clear section delineation with descriptive headers
- Context-sensitive help text for technical terms
- Error prevention mechanisms including response validation
- Intuitive navigation between sections

Pilot Testing and Expert Review

The technical implementation was validated through two phases:

1. Initial functionality testing to verify LimeSurvey’s operational status
2. Pilot testing with nine experts from relevant medical and non-medical fields, as shown in Figure 1, who assessed both usability and technical performance

Step 3. Pilot testing Study

A pilot study was conducted on 206 responders (203 completed questionnaires analyzed), to assess the questionnaire’s ability to capture meaningful variations between different groups of physicians and to identify potential improvements needed. Table 2 reports on the characteristics of the participants. The average age was 41 (SD=14), approximately 51% were residents, and 43% were identified as men.

**Table 2.** - Participant characteristics (n=203) -.

| Characteristics | N (%) or mean (SD) |
| --- | --- |
| Sex |  |
| Male | 88 (43.3%) |
| Female | 113 (55.7%) |
| Other | 2 (1.0%) |
| Age | 41.2 (14.1) |
| Specialization status |  |
| Residents | 103 (50.7%) |
| Specialists | 100 (49.3%) |
| Years workers | 12.4 (13.2) |
| Did you have any specific training in digital or information technologies during your studies? |  |
| Yes | 35 (17%) |
| No | 168 (83%) |

The questionnaire’s ability to differentiate knowledge levels and detect variations between professional groups was assessed through the analysis of responses to the AI knowledge section, as shown in Table 3. The results demonstrate the instrument’s sensitivity in capturing different levels of AI knowledge and experience.

The study evidenced that 17% of the sample did have specific training in digital or information technologies during their medical course, 22% attended courses or seminars on AI in medicine, and 21% declared “no knowledge of AI”. Among the AI applications, the sample reported that Knowledge in diagnostics was higher (49%), and there were no significant differences between groups (residents versus specialists), as reported in Table 3.

**Table 3.** - Knowledge about AI Technology in Medicine -.

|  | **Specialization status** |  |  |  |
| --- | --- | --- | --- | --- |
|  | **Residents**<br>**(N=103)** | **Specialists**<br>**(N=100)** | **Total** | **p** |
|  | N (%) | N (%) | 203 (100.0%) |  |
| How would you describe your general knowledge of AI? |  |  |  |  |
| None | 19 (18.4%) | 25 (25.0%) | 44 (21.7%) | 0.310 |
| Basic knowledge | 70 (68.0%) | 60 (60.0%) | 130 (64.0%) |  |
| Intermediate knowledge | 14 (13.6%) | 13 (13.0%) | 27 (13.3%) |  |
| Advanced knowledge | 0 (0.0%) | 2 (2.0%) | 2 (1.0%) |  |
| Have you attended any courses or seminars on AI in medicine? |  |  |  |  |
| No | 84 (81.6%) | 82 (82.0%) | 166 (81.8%) | 0.934 |
| Yes | 19 (18.4%) | 18 (18.0%) | 37 (18.2%) |  |
| Which of the following AI applications do you know? |  |  |  |  |
| Diagnostic imaging (radiology, dermatology) |  |  |  |  |
| No | 57 (55.3%) | 47 (47.0%) | 104 (51.2%) | 0.235 |
| Yes | 46 (44.7%) | 53 (53.0%) | 99 (48.8%) |  |
| Clinical decision support systems |  |  |  |  |
| No | 79 (76.7%) | 74 (74.0%) | 153 (75.4%) | 0.655 |
| Yes | 24 (23.3%) | 26 (26.0%) | 50 (24.6%) |  |
| Care planning and management |  |  |  |  |
| No | 87 (84.5%) | 91 (91.0%) | 178 (87.7%) | 0.157 |
| Yes | 16 (15.5%) | 9 (9.0%) | 25 (12.3%) |  |
| Patient data monitoring and analysis |  |  |  |  |
| No | 69 (67.0%) | 76 (76.0%) | 145 (71.4%) | 0.155 |
| Yes | 34 (33.0%) | 24 (24.0%) | 58 (28.6%) |  |
| Interpretation of reports (ECG, EEG, etc.) |  |  |  |  |
| No | 62 (60.2%) | 63 (63.0%) | 125 (61.6%) | 0.681 |
| Yes | 41 (39.8%) | 37 (37.0%) | 78 (38.4%) |  |
| Machine learning in diagnosis |  |  |  |  |
| No | 85 (82.5%) | 89 (89.0%) | 174 (85.7%) | 0.187 |
| Yes | 18 (17.5%) | 11 (11.0%) | 29 (14.3%) |  |
| Machine learning and Big Data for drug development |  |  |  |  |
| No | 90 (87.4%) | 88 (88.0%) | 178 (87.7%) | 0.893 |
| Yes | 13 (12.6%) | 12 (12.0%) | 25 (12.3%) |  |
| Virtual nursing assistants |  |  |  |  |
| No | 98 (95.1%) | 99 (99.0%) | 197 (97.0%) | 0.105 |
| Yes | 5 (4.9%) | 1 (1.0%) | 6 (3.0%) |  |
| Chatbot for diagnostic queries |  |  |  |  |
| No | 73 (70.9%) | 75 (75.0%) | 148 (72.9%) | 0.508 |
| Yes | 30 (29.1%) | 25 (25.0%) | 55 (27.1%) |  |
| Monitoring, home care and predictive analytics |  |  |  |  |
| No | 95 (92.2%) | 94 (94.0%) | 189 (93.1%) | 0.619 |
| Yes | 8 (7.8%) | 6 (6.0%) | 14 (6.9%) |  |
| Personalized medicine |  |  |  |  |
| No | 92 (89.3%) | 89 (89.0%) | 181 (89.2%) | 0.941 |
| Yes | 11 (10.7%) | 11 (11.0%) | 22 (10.8%) |  |
| Improving genome editing |  |  |  |  |
| No | 99 (96.1%) | 97 (97.0%) | 196 (96.6%) | 0.730 |
| Yes | 4 (3.9%) | 3 (3.0%) | 7 (3.4%) |  |
| Electronic health records (EHR) and data analytics |  |  |  |  |
| No | 76 (73.8%) | 70 (70.0%) | 146 (71.9%) | 0.548 |
| Yes | 27 (26.2%) | 30 (30.0%) | 57 (28.1%) |  |
| Robotic surgery procedures AI-based |  |  |  |  |
| No | 89 (86.4%) | 85 (85.0%) | 174 (85.7%) | 0.774 |
| Yes | 14 (13.6%) | 15 (15.0%) | 29 (14.3%) |  |
| None |  |  |  |  |
| No | 78 (75.7%) | 74 (74.0%) | 152 (74.9%) | 0.777 |
| Yes | 25 (24.3%) | 26 (26.0%) | 51 (25.1%) |  |

Overall, only 19.2% of participants reported using AI tools in clinical practice, with no significant differences between residents (17.5%) and specialists (21.0%) (p = 0.524). The most used applications were diagnostic imaging (6.9%), chatbots for diagnostic queries (8.4%), and clinical decision support systems (5.4%). Other applications, such as machine learning in diagnosis (2.5%) and electronic health record analytics (3.4%), had very low adoption rates. At the same time, tools like drug discovery, genome editing, and robotic surgery were nearly unused. No significant differences were observed between the groups for all applications (all p > 0.05), indicating a generally low level of AI integration into clinical practice (Table 4).

**Table 4.**
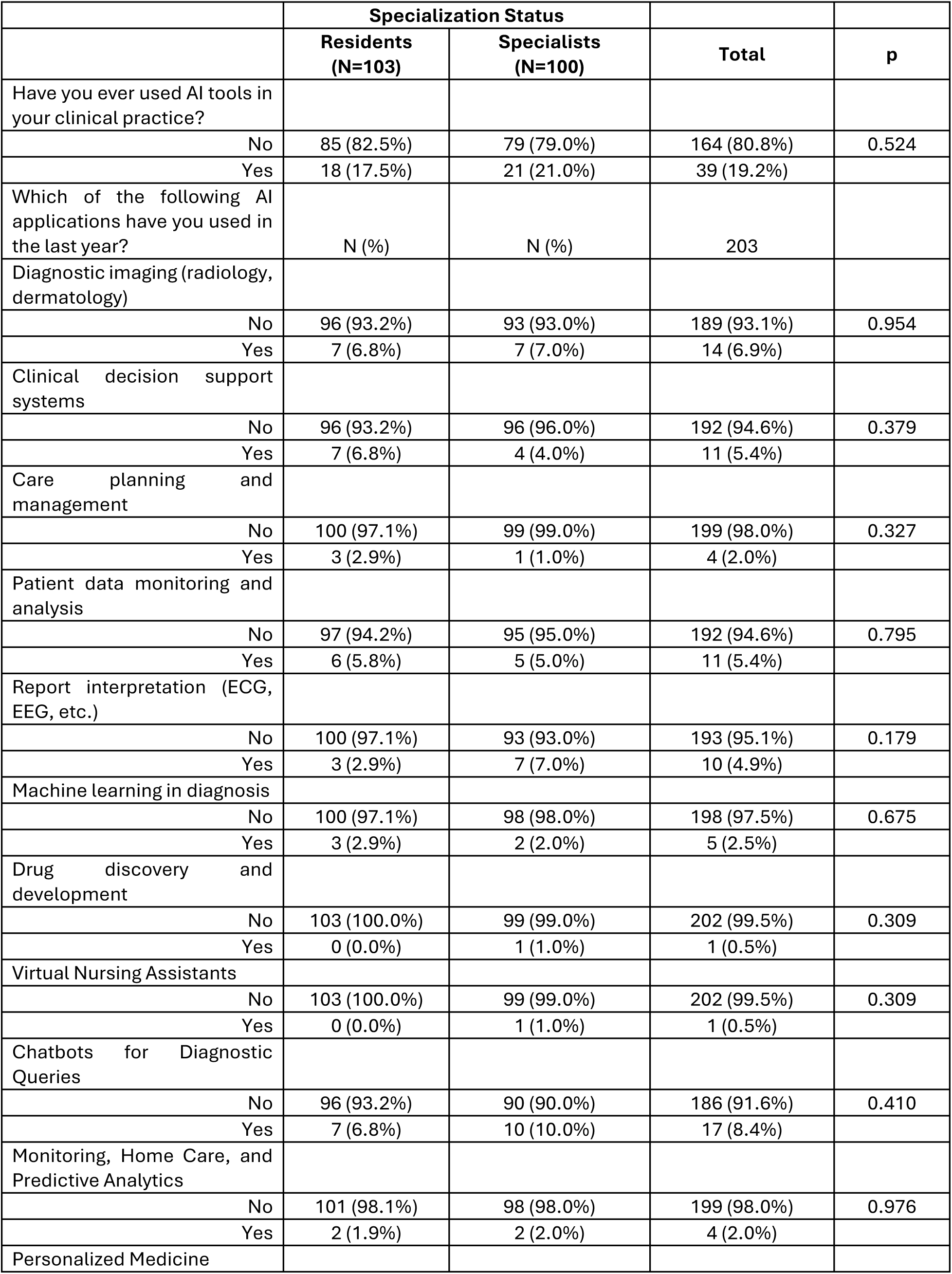
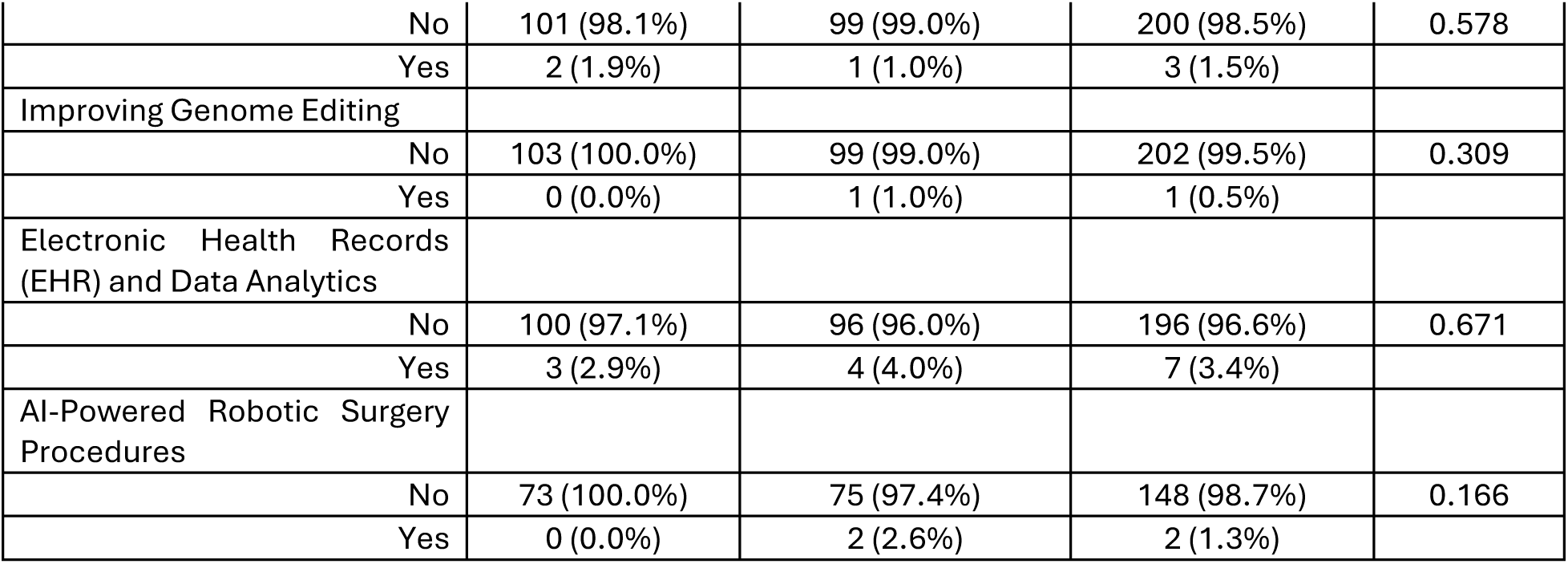
- AI Use and Experience in Medicine in the last year -.

Table 5 displays the results of the instrument’s effectiveness evaluation in measuring attitudes and awareness using a five-point Likert scale. Participants demonstrated consistently positive attitudes toward AI’s role in medicine, with high mean scores across groups for its potential in differential diagnosis (mean 4.9, SD 0.6) and improving therapeutic prescriptions (mean 4.6, SD 0.8). No significant differences were observed between residents and specialized participants in any category (p > 0.05). The strong consensus was on the need for additional training in AI for doctors (mean 5.4, SD 0.5), highlighting a key area for curriculum development. Concerns about AI posing a threat to the doctor’s role were moderate (mean 3.4, SD 0.9).

**Table 5.**
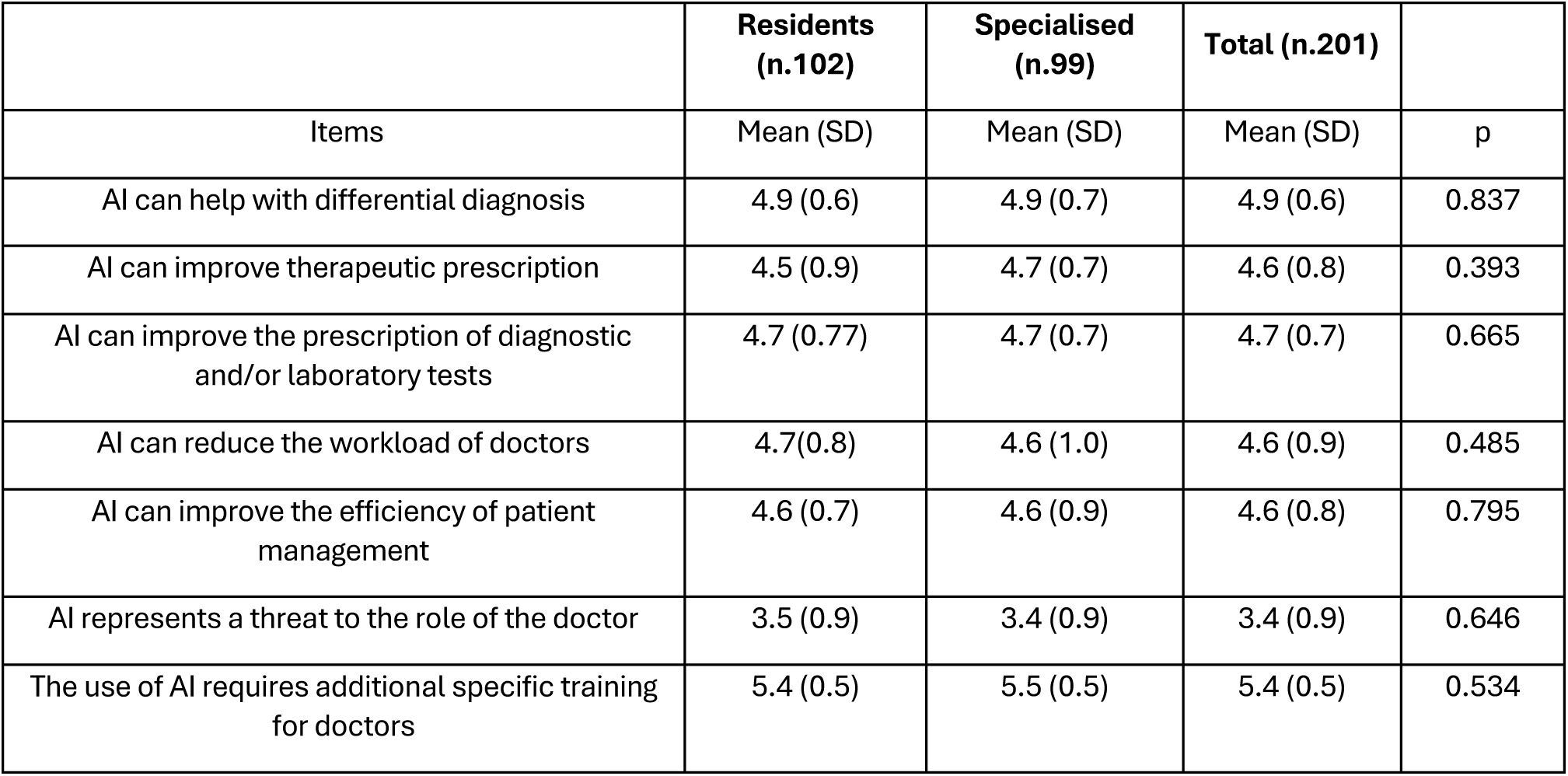
- Attitudes and Awareness Regarding AI in Medicine -.

The analysis presented in Table 6 evaluated the questionnaire’s capacity to assess willingness to adopt AI and identify barriers and facilitators. Most respondents expressed a positive attitude toward integrating AI into clinical practice, with 26.6% indicating they were “very available” and 53.2% stating they were “available”. There were no significant differences observed between residents and specialists (p=0.757). The primary barriers identified included a lack of training (80.3%) and resistance to change (53.7%), with no significant differences between the two groups. Additionally, 38.4% of participants voiced concerns about privacy and data security. Residents were more likely to report a lack of scientific evidence regarding the effectiveness (34.0%) compared to specialists (21.0%) (p=0.039). Education and professional development (87.7%) were the most endorsed incentives for integrating AI and ongoing technical support (50.2%). Notably, residents emphasized the need for continuous technical support more than specialists, with 58.3% of residents versus 42.0% of specialists expressing this view (p=0.021). Furthermore, 54.2% of participants highlighted the importance of scientific evidence regarding efficacy and safety, with a higher percentage of residents (61.2%) compared to specialists (47.0%) recognizing this need (p=0.043).

**Table 6.**
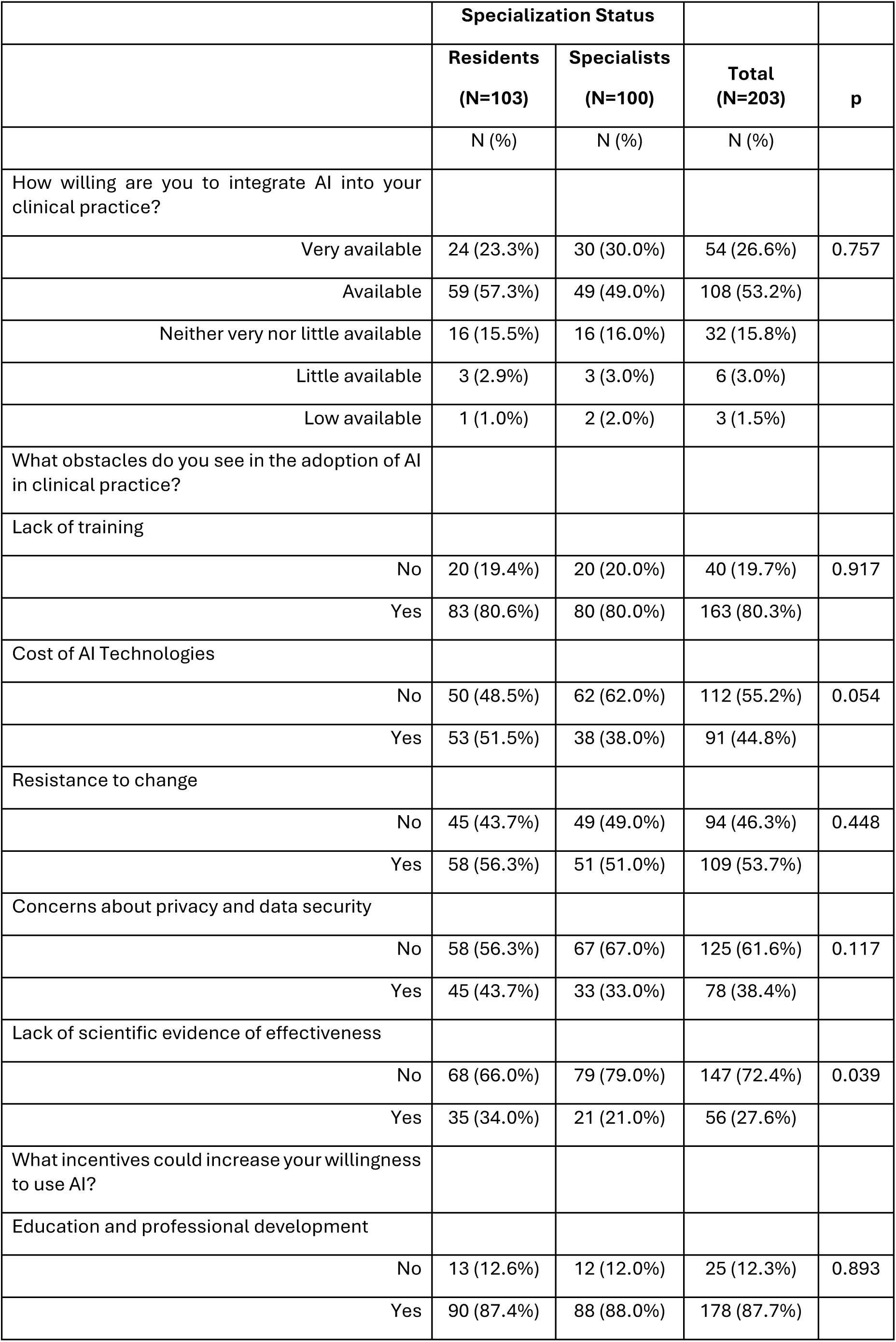
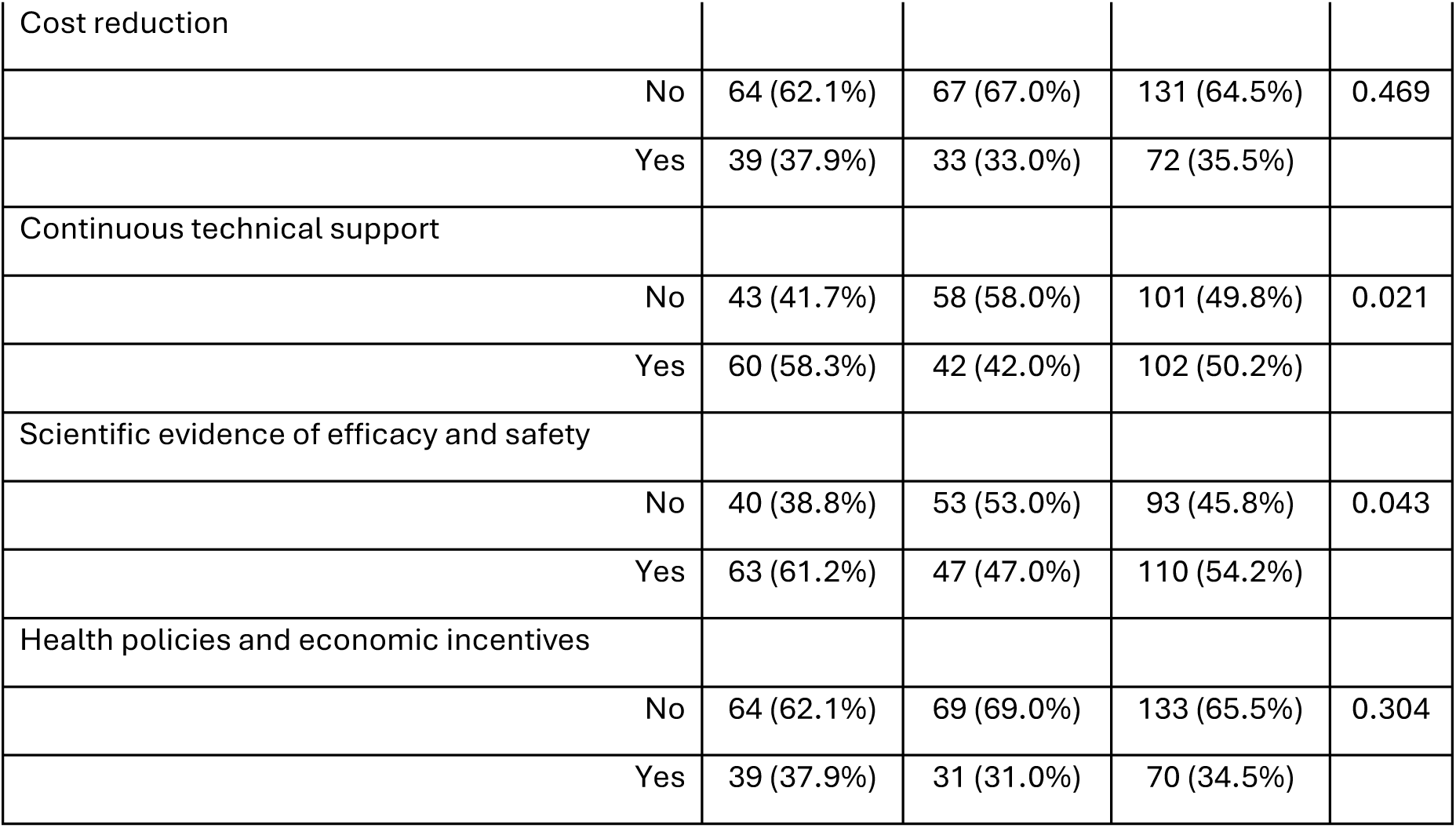
- Willingness to Use AI in Medicine –.

|  | **Specialization Status** |  |  |  |
| --- | --- | --- | --- | --- |
|  | **Residents**<br>**(N=103)** | **Specialists**<br>**(N=100)** | **Total**<br>**(N=203)** | **p** |
|  | N (%) | N (%) | N (%) |  |
| How willing are you to integrate AI into your clinical practice? |  |  |  |  |
| Very available | 24 (23.3%) | 30 (30.0%) | 54 (26.6%) | 0.757 |
| Available | 59 (57.3%) | 49 (49.0%) | 108 (53.2%) |  |
| Neither very nor little available | 16 (15.5%) | 16 (16.0%) | 32 (15.8%) |  |
| Little available | 3 (2.9%) | 3 (3.0%) | 6 (3.0%) |  |
| Low available | 1 (1.0%) | 2 (2.0%) | 3 (1.5%) |  |
| What obstacles do you see in the adoption of AI in clinical practice? |  |  |  |  |
| Lack of training |  |  |  |  |
| No | 20 (19.4%) | 20 (20.0%) | 40 (19.7%) | 0.917 |
| Yes | 83 (80.6%) | 80 (80.0%) | 163 (80.3%) |  |
| Cost of AI Technologies |  |  |  |  |
| No | 50 (48.5%) | 62 (62.0%) | 112 (55.2%) | 0.054 |
| Yes | 53 (51.5%) | 38 (38.0%) | 91 (44.8%) |  |
| Resistance to change |  |  |  |  |
| No | 45 (43.7%) | 49 (49.0%) | 94 (46.3%) | 0.448 |
| Yes | 58 (56.3%) | 51 (51.0%) | 109 (53.7%) |  |
| Concerns about privacy and data security |  |  |  |  |
| No | 58 (56.3%) | 67 (67.0%) | 125 (61.6%) | 0.117 |
| Yes | 45 (43.7%) | 33 (33.0%) | 78 (38.4%) |  |
| Lack of scientific evidence of effectiveness |  |  |  |  |
| No | 68 (66.0%) | 79 (79.0%) | 147 (72.4%) | 0.039 |
| Yes | 35 (34.0%) | 21 (21.0%) | 56 (27.6%) |  |
| What incentives could increase your willingness to use AI? |  |  |  |  |
| Education and professional development |  |  |  |  |
| No | 13 (12.6%) | 12 (12.0%) | 25 (12.3%) | 0.893 |
| Yes | 90 (87.4%) | 88 (88.0%) | 178 (87.7%) |  |
| Cost reduction |  |  |  |  |
| No | 64 (62.1%) | 67 (67.0%) | 131 (64.5%) | 0.469 |
| Yes | 39 (37.9%) | 33 (33.0%) | 72 (35.5%) |  |
| Continuous technical support |  |  |  |  |
| No | 43 (41.7%) | 58 (58.0%) | 101 (49.8%) | 0.021 |
| Yes | 60 (58.3%) | 42 (42.0%) | 102 (50.2%) |  |
| Scientific evidence of efficacy and safety |  |  |  |  |
| No | 40 (38.8%) | 53 (53.0%) | 93 (45.8%) | 0.043 |
| Yes | 63 (61.2%) | 47 (47.0%) | 110 (54.2%) |  |
| Health policies and economic incentives |  |  |  |  |
| No | 64 (62.1%) | 69 (69.0%) | 133 (65.5%) | 0.304 |
| Yes | 39 (37.9%) | 31 (31.0%) | 70 (34.5%) |  |

Assessing willingness to engage in artificial intelligence medical training, the analysis demonstrated overwhelming interest in AI education, with 95.1% of respondents expressing willingness to participate in training programs. Both career stages showed a consistent level of interest, with 93.2% of residents and 97.0% of specialists indicating interest, and there were no significant differences (p=0.211).

Regarding preferred training modalities, online courses emerged as the most popular format, with 61.1% of all respondents favouring this option. Residents showed a somewhat higher preference for online learning (67.0%) compared to specialists (55.0%), though this difference did not reach statistical significance (p=0.080). The second most preferred modality was hands-on training, with 60.6% of respondents choosing it, with almost identical preferences among residents (60.2%) and specialists (61%), p=0.906.

In-person seminars and workshops were preferred by 45.3% of respondents, with specialists showing greater interest (51.0%) than residents (39.8%; p=0.109). Webinars and conferences were the least preferred format, with only 29.6% of respondents expressing interest in this modality. This preference was consistent across both groups, with 29.1% of residents and 30.0% of specialists selecting this option (p=0.892).

The absence of statistically significant differences in preferences between residents and specialists suggests that career stage may not determining the choice of AI learning formats.

Universally Applicable Scenario

The instrument’s ability to measure clinical decision-making alignment with AI was assessed by validating clinical scenarios.

The universal scenario was structured as follows:

“A 32-year-old Caucasian patient has reported asthenia and evening fever for about 10 days. In medical history: cocaine addiction, history of stage IV Hodgkin lymphoma treated with chemotherapy in complete remission at the last follow-up in 2023, unprotected sexual intercourse, previous HPV infection. During the physical exam, it was determined that there were inguinal lymphadenopathy and a single genital ulcerative lesion in the balano-preputial area”.

For this scenario, participants were asked to rate their feelings towards three diagnoses: the first AI-generated diagnosis and two other AI proposals generated for other clinical scenarios

In the general scenario, clinical agreement with the diagnosis among those proposed by AI was 91% (among specialist doctors: 92%), as reported in Table 8. The consistent response patterns across different professional groups further support the reliability of this section of the questionnaire.

**Table 7.** - Willingness to Learn AI in Medicine -.

|  | Specialization Status |  |  |  |
| --- | --- | --- | --- | --- |
|  | Residents<br>(N=103) | Specialists<br>(N=100) | Total<br>(N=203) | p |
|  | N (%) | N (%) | N (%) |  |
| Would you be interested in participating in training programs on AI applied to medicine? |  |  |  |  |
| No | 7 (6.8%) | 3 (3.0%) | 10 (4.9%) | 0.211 |
| Yes | 96 (93.2%) | 97 (97.0%) | 193 (95.1%) |  |
| Which modalities would you prefer? |  |  |  |  |
| Online courses |  |  |  |  |
| No | 34 (33.0%) | 45 (45.0%) | 79 (38.9%) | 0.080 |
| Yes | 69 (67.0%) | 55 (55.0%) | 124 (61.1%) |  |
| In-person seminars and workshops |  |  |  |  |
| No | 62 (60.2%) | 49 (49.0%) | 111 (54.7%) | 0.109 |
| Yes | 41 (39.8%) | 51 (51.0%) | 92 (45.3%) |  |
| Hands-on training |  |  |  |  |
| No | 41 (39.8%) | 39 (39.0%) | 80 (39.4%) | 0.906 |
| Yes | 62 (60.2%) | 61 (61.0%) | 123 (60.6%) |  |
| Webinars and conferences |  |  |  |  |
| No | 73 (70.9%) | 70 (70.0%) | 143 (70.4%) | 0.892 |
| Yes | 30 (29.1%) | 30 (30.0%) | 60 (29.6%) |  |

**Table 8.** - Agreement between Physicians and AI -.

|  | Specialization Status |  |  |  |
| --- | --- | --- | --- | --- |
|  | Residents<br>(N=103) | Specialists<br>(N=100) | Total (N=203) | p |
|  | N (%) | N (%) | N (%) |  |
| **Differential diagnosis:** |  |  |  |  |
| Syphilis |  |  |  |  |
| Strongly disagree | 2 (1.9%) | 2 (2.0%) | 4 (2.0%) | 0.902 |
| Disagree | 1 (1.0%) | 0 (0.0%) | 1 (0.5%) |  |
| Neutral | 7 (6.8%) | 6 (6.0%) | 13 (6.4%) |  |
| Agree | 68 (66.0%) | 68 (68.0%) | 136 (67.0%) |  |
| Strongly agree | 25 (24.3%) | 24 (24.0%) | 49 (24.1%) |  |
| Irritable Bowel Syndrome (IBS) |  |  |  |  |
| Strongly disagree | 37 (35.9%) | 31 (31.0%) | 68 (33.5%) | 0.823 |
| Disagree | 52 (50.5%) | 54 (54.0%) | 106 (52.2%) |  |
| Neutral | 11 (10.7%) | 11 (11.0%) | 22 (10.8%) |  |
| Agree | 3 (2.9%) | 3 (3.0%) | 6 (3.0%) |  |
| Strongly agree | 0 (0.0%) | 1 (1.0%) | 1 (0.5%) |  |
| Pathology inflammatory pelvic |  |  |  |  |
| Strongly disagree | 15 (14.6%) | 13 (13.0%) | 28 (13.8%) | 0.798 |
| Disagree | 36 (35.0%) | 36 (36.0%) | 72 (35.5%) |  |
| Neutral | 29 (28.2%) | 24 (24.0%) | 53 (26.1%) |  |
| Agree | 22 (21.4%) | 24 (24.0%) | 46 (22.7%) |  |
| Strongly agree | 1 (1.0%) | 3 (3.0%) | 4 (2.0%) |  |

Step 4: Internal Consistency

The reliability analysis of the 8-item scale (opinion and willingness to use AI), yielded a Cronbach’s Alpha of α=0.7481, indicating acceptable internal consistency as reported in Table 9. Item-level statistics showed that most items had moderate to high item-test correlations (Item 1= 0.7474; Item 8 = 0.7178). However, two items, Item 6 and Item 7, had lower item-test correlations (0.3277 and 0.3713, respectively), suggesting weaker alignment with the overall construct.

**Table 9.** - Reliability statistics –.

|  | Average |  |  |  |  |  |
| --- | --- | --- | --- | --- | --- | --- |
|  | N | Sign correlation | Item test correlation | Item-rest | interitem | Alpha |
| **Opinion and willing to use AI** |  |  |  |  |  |  |
| ITEM 1. AI can help with differential diagnosis | 203 | + | 0.7474 | 0.6305 | 0.2373 | 0.6853 |
| ITEM 2. AI can improve therapeutic prescription | 203 | + | 0.7219 | 0.5963 | 0.2431 | 0.6922 |
| ITEM 3. AI can improve the prescription of diagnostic and/or laboratory tests | 203 | + | 0.7126 | 0.5840 | 0.2453 | 0.6947 |
| ITEM 4. AI can reduce the workload of doctors | 203 | + | 0.5548 | 0.3850 | 0.2814 | 0.7327 |
| ITEM 5. AI can improve the efficiency of patient management | 203 | + | 0.6591 | 0.5145 | 0.2575 | 0.7083 |
| ITEM 6.AI represents a threat to the role of the doctor | 203 | - | 0.3277 | 0.1259 | 0.3335 | 0.7779 |
| ITEM 7. The use of AI requires additional specific training for doctors | 203 | + | 0.3713 | 0.1734 | 0.3235 | 0.7700 |
| ITEM 8. Willing to integrate AI into your clinical practice | 203 | - | 0.7178 | 0.5909 | 0.2441 | 0.6933 |
| **Test scale** |  |  |  |  | **0.2707** | **0.7481** |

The average interim correlation was 0.2707, within the acceptable range of measures assessing a unidimensional construct.

The interim correlation matrix (Figure 2) revealed moderate to strong correlations among most items (Item 1 and Item 2 = 0.58). In contrast, certain items, such as Item 6, exhibited weaker correlations with other items (Item 6 and Item 3 = 0.03).

Excluding specific items such as Item 6 and Item 7 led to an increase in Cronbach’s Alpha (0.7779 and 0.7700, respectively), suggesting that these items may contribute less to the scale’s overall reliability.

The internal consistency of the dichotomous items was assessed using Kuder-Richardson Formula 21 (KR-21), which is calculated as follows (Kuder & Richardson, 1937):

KR-21=k/(k-1)(1-((M·(k-M))/(k ·var))

Where:

- k=18 (total number of items),
- M=10 (mean of total scores),
- var. =20.7044 (variance of total scores).

Using this formula, the KR-21 coefficient was calculated to be ρ=0.832.

The final version of the Italian Knowledge, Attitudes, Practice, and Clinical Agreement between Medical Doctors and Artificial Intelligence Questionnaire (I-KAPCAM-AI-Q) questionnaire was reported in supplementary material.

## Discussion

The validation of the I-KAPCAM-AI-Q marks a crucial advancement in comprehending and enhancing the integration of AI within Italian healthcare system. This aligns with the recommendations of the Italian National Health Care System for utilizing digital tools in delivering healthcare. A tool like the I-KAPCAM-AI-Q contributes to informing policy development regarding AI integration in healthcare settings, as well as guiding institutional training programs and educational initiatives, and supporting strategic planning for AI implementation in clinical practice. Moreover, it is useful in helping to track changes in attitudes and practices over time. Our findings reveal several key insights that warrant detailed discussion. The I-KAPCAM-AI-Q demonstrated robust psychometric properties, with strong content validity (S-CVI/Ave=0.98) and acceptable internal consistency (Cronbach’s Alpha=0.7481). The KR-21 coefficient of 0.832 for dichotomous items further supports the instrument’s reliability. These metrics align with or exceed those of similar validated healthcare questionnaires, suggesting that the I-KAPCAM-AI-Q is a reliable tool for assessing AI readiness in healthcare settings. For such a reason, I-KAPCAM-AI-Q holds potential for use in experimental studies related to AI, such as longitudinal studies about the adoption of AI in healthcare. It allows for comparative analysis across different medical specialities and supports evidence-based approaches to AI implementation. While the data collected in this pilot study cannot be generalized due to the study’s specific objective, they nonetheless highlight interesting findings that can be further developed through targeted research. The I-KAPCAM-AI-Q builds on previous research assessing AI literacy and attitudes in healthcare but introduces significant methodological advancements. A striking finding from our preliminary study is the substantial growth in digital technology training, with only 17% of participants reporting specific training during their medical education. This deficit is particularly concerning given the rapid advancement of AI in healthcare, and aligns with findings from the VALIDATE Project regarding insufficient digital health preparation among Italian medical professionals. The lack of formal training may explain why only 19% of respondents reported using AI applications in their practice, despite high interest and willingness to integrate AI tools. Similar findings were reported by Mousavi Baigi et al. [27], who found that only 15-22% of healthcare students received formal AI training across multiple European countries. The low AI training levels in our sample (17%) further confirm that the current medical education system does not adequately prepare physicians for AI-based healthcare.

The comparison between residents and specialists revealed interesting patterns. While both groups showed similar levels of AI knowledge and general attitudes, residents demonstrated significantly higher interest in technical support (58.3% vs 42.0%, p=0.021) and evidence-based validation (61.2% vs 47.0%, p=0.043). This difference suggests that newer medical professionals may be more attuned to the importance of systematic implementation and validation of AI tools, possibly reflecting evolving perspectives in medical education. AlZaabi et al., [28] reported similar generational differences in AI readiness among physicians, with younger practitioners showing greater technological adaptability. The use of AI in medical practice reveals distinct patterns between junior doctors and specialists, highlighting crucial considerations for medical education and implementation [50–53]. Junior doctors typically demonstrate greater capacity to embrace new technologies, but possess less clinical experience to contextualise AI outputs, potentially leading to over-reliance on AI recommendations without the benefit of extensive clinical intuition. Specialists, drawing from years of practice, have developed robust mental models and decision-making frameworks that enable them to evaluate AI applications critically. While their established workflows may create initial resistance to AI adoption, their deep specialty knowledge allows them to precisely identify where AI adds value versus where it might complicate existing processes [54–55]. Generational differences in AI adoption observed in our study highlight the evolving perspective of younger physicians.

The results indicate a preference for flexible learning options (online courses) and practical experience (hands-on training) over traditional educational formats. The rapid expansion of medical knowledge makes it difficult for individual practitioners to stay fully informed about all advancements [34]. Our findings The analysis demonstrates concerning levels of trust in ChatGPT for medical diagnostics, with 8.4% of participants regularly using the chatbot for diagnostic inquiries. The proportion of participants using ChatGPT for diagnostic inquiries (8.4%) justifies our methodological decision to investigate the agreement between ChatGPT and participant responses regarding real-life medical scenarios. Despite its limitations, this finding has real-world clinical relevance, as healthcare consumers are already turning to ChatGPT for medical decision-making [56]. The high clinical agreement (91%) with AI-proposed diagnoses in the universal scenario is noteworthy, suggesting that healthcare providers can effectively evaluate AI-generated clinical recommendations, despite limited formal training. This finding has important implications for future integration of AI in clinical decision support systems and highlights the potential for AI to complement rather than replace clinical judgment.

The validation process of the I-KAPCAM-AI-Q has two main methodological limitations: the lack of a test-retest reliability assessment and the potential selection bias stemming from our volunteer-based sampling approach. Although these limitations are specific to the validation methodology, the pilot study conducted as part of this process has yielded valuable insights that will inform future applications of the tool. Future applications of the validated I-KAPCAM-AI-Q could reveal multiple important research priorities. While our pilot study was instrumental in refining and validating the questionnaire, its findings should be viewed primarily as supportive of the tool’s development rather than as generalizable results. Future large-scale applications of the I-KAPCAM-AI-Q should focus on conducting longitudinal studies to evaluate how knowledge and attitudes of physicians towards AI evolve over time with increased exposure to AI technologies. The questionnaire could be implemented with more diverse clinical scenarios across different medical specialties, to better understand the tool’s effectiveness in various healthcare contexts. Future research through the I-KAPCAM-AI-Q can investigate variations in responses across different healthcare settings, geographical regions, and levels of technological infrastructure and the relationship between measured AI readiness and actual implementation success in clinical settings. Furthermore, future research should explore how the I-KAPCAM-AI-Q can be adapted to assess AI readiness in different healthcare systems and cultural contexts while maintaining its psychometric properties. The tool’s comprehensive assessment of technical competency and clinical judgment makes it particularly suitable for developing targeted educational interventions across Italian healthcare, from university-level training programs to workplace implementation strategies. Several distinctive strengths enhance the significance of the I-KAPCAM-AI-Q in the evolving landscape of AI assessment tools. A primary strength lies in its demonstrated effectiveness across different levels of medical expertise, with particular value for early-career physicians and residents. The evaluation of AI readiness among young doctors is especially crucial as they represent the future of healthcare and are often at the forefront of technological adoption. Notably, the I-KAPCAM-AI-Q stands as the first and only Italian validated instrument that systematically compares clinical decision-making with ChatGPT responses using real-world scenarios. This unique feature provides unprecedented insights into the alignment between human and artificial intelligence, reasoning in authentic clinical situations. The use of real clinical cases rather than theoretical scenarios enhances the tool’s practical relevance and validity.

The tool’s power lies in its ability to assess clinical agreement, not only between physicians and AI, but also among healthcare providers themselves. This capability opens valuable avenues for understanding how different clinicians approach similar cases and how their decision-making patterns align or diverge when using AI tools. These perspectives are essential for developing standardized approaches to integrating AI in clinical practice and identifying areas where additional training or support may be required. These findings, combined with the tool’s robust psychometric properties, ensure that I-KAPCAM-AI-Q is a comprehensive and innovative framework for advancing AI education and implementation in medicine. The ability to simultaneously assess technical competency, clinical judgment, and agreement patterns makes it an essential instrument for guiding the future of AI integration in healthcare settings.

## Conclusions

The I-KAPCAM-AI-Q provides a strong foundation for assessing AI readiness in healthcare. Its validation, supported by expert review and pilot testing, establishes its reliability and relevance. This tool informs the development of targeted educational programs by identifying knowledge gaps and assessing attitudes toward AI. High clinical agreement with AI diagnoses underscores the potential for AI to complement clinical judgment. The I-KAPCAM-AI-Q is a valuable resource for advancing AI education and integration, shaping the future of healthcare in the era of artificial intelligence.

## Data Availability

All data produced in the present study are available upon reasonable request to the authors

## Supplementary Materials

The following supporting information can be downloaded at: www.mdpi.com/xxx/s1.

## Author Contributions

“Conceptualization, V.C. and E.B.; methodology, V.C., S.N.; software, E.B.; validation, V.C; E.B; L.P.; G.D.P.; E.C.; M.Ma.; M.Mu.; E.P.; G.P. L.T.; A.B.; G.D.; M.G.; F.M.; L.F.; S.N.; formal analysis, V.C.; investigation, V.C; L.P.; E.B; G.D.P.; E.C.; M.Ma.; M.Mu.; E.P.; G.P.; P.P.; L.T.; A.B.; G.D.; M.G.; F.M.; L.F.; S.N. data curation, V.C. and E.B.; writing—original draft preparation, V.C.; E.B.; L.P.; M.Ma.; writing—review and editing, V.C.; E.B.; L.P.; G.D.P.; M.Ma.; M.Mu; S.N.; supervision, V.C. All authors have read and agreed to the published version of the manuscript.

## Informed Consent Statement

Informed consent was obtained from all subjects involved in the study.

## Data Availability Statement

The raw data supporting the conclusions of this article will be made available by the authors upon request.

## Acknowledgments

Authors thank all participants to all activity in the current study

